# Nanomechanical and molecular characterization of aging in dentinal collagen

**DOI:** 10.1101/2021.08.31.21262730

**Authors:** Christina MAP Schuh, Camila Leiva-Sabadini, Sophia Huang, Nelson P Barrera, Laurent Bozec, Sebastian Aguayo

## Abstract

Methylglyoxal (MGO) is an important molecule derived from glucose metabolism with the capacity of attaching to collagen and generating advanced glycation end-products (AGEs), which accumulate in tissues over time and are associated to aging and diseases. However, the accumulation of MGO-derived AGEs in dentin and their effect on the nanomechanical properties of dentinal collagen remain unknown. Thus, the aim of the present study was to quantify MGO-based AGEs in the organic matrix of human dentin as a function of age and associate these changes with alterations in the nanomechanical and ultrastructural properties of dentinal collagen. For this, twelve healthy teeth from <26 year-old and >50 year-old patients were collected and prepared to obtain crown and root dentin discs. Following demineralization, MGO-derived AGEs were quantified with a competitive ELISA assay. In addition, atomic force microscopy (AFM) nano-indentation was utilized to measure changes in elastic modulus in both peritubular (PT) and intertubular (IT) collagen fibrils. Finally, principal component analysis (PCA) was carried out to determine aging profiles for both crown and root dentin. Results showed increased presence of MGO AGEs in the organic matrix of dentin in the >50 year-old compared to <26 year-old samples in both crown and root. Furthermore, an overall increase in PT and IT collagen elasticity was observed in the >50 year-old group associated to ultrastructural changes in the organic matrix determined by AFM analysis. Furthermore, PCA loading plots suggested different ‘aging profiles’ in both crown and root dentin, which could potentially have important therapeutic implications in restorative and adhesive dentistry approaches. Overall, these results demonstrate that the organic matrix of human dentin undergoes aging-related changes due to MGO-derived AGEs with important changes in nanomechanical behavior of collagen that may impact diagnostic and restorative procedures in the elderly.

## Introduction

In recent years, populations worldwide have experienced a rapid increase in life expectancy that has brought forward many challenges in diagnostics and treatment for older people (Prince et al. 2015). Elderly people are also likely to suffer from important oral pathologies such as periodontal disease, dental caries (especially root caries) and oral candidiasis (Murray Thomson 2014). As the process of aging is complex and involves an important number of biological and molecular changes in cells, tissues, and organs (DiLoreto and Murphy 2015; Sui et al. 2016), it remains essential to comprehend its impact on the pathogenesis of relevant oral diseases and restorative approaches.

Recently, strong attention has been placed on investigating age-related changes in collagen (Gurav 2013; Ahmed et al. 2017), since they are the most abundant structural proteins in the body (Capella-Monsonís et al. 2018) and one of the main constituents of the organic matrix of dentin and periodontal tissues (Goldberg 2011). In modern restorative dentistry, dentinal collagen has become an important substrate for resin-dentin bonding and has been associated to the success or failure of the hybrid layer (Breschi et al. 2018). One of the most important age-associated changes in collagen results from the slow and irreversible accumulation of advanced glycation end-products (AGEs) (Nass et al. 2007). AGEs form sugar-residue crosslinks within the collagen and are often considered to have deleterious effects on the biophysical and biochemical properties of collagen-rich tissues. Some of the most medically relevant AGEs include glucosepane, pentosidine, and methyl-glyoxal (MGO)-derived AGEs (Gkogkolou and Böhm 2012). MGO is derived from glucose metabolism and has the ability of attaching to lysine and arginine residues on the collagen molecule (Fessel et al. 2014; Wetzels et al. 2017). This process drives the formation of AGE-mediated adducts and crosslinks in a time-dependent manner; thus, the accumulation of AGEs is higher in tissues with slow collagen turnaround and in aged tissues (Ahmed et al. 2017). AGEs have been implied in a wide range of chronic diseases, and AGE-mediated protein modification is believed to be responsible for increasing the stiffness and fragility of collagen in human skin and bone (Snedeker and Gautieri 2014; Poundarik et al. 2015). To date, some studies have explored the effect of aging on the accumulation of certain AGEs in dentin, such as pentosidine (Shinno et al. 2016; Greis et al. 2018); and at the macroscale, it has been shown that AGE accumulation in dentin results in altered mechanical properties including higher tendencies to fracture under stress. However, not much is known regarding the accumulation of MGO-derived AGEs in dentin, or on the effect that these changes could generate on the ultrastructural and mechanical properties of dentinal collagen at the nanoscale level.

In recent years, major advances in the field of atomic force microscopy (AFM) have potentiated the non-destructive study of biological samples, by combining topographical and nanomechanical assessment with nanoscale resolution (Dufrêne et al. 2021). Utilizing AFM, it is possible to not only characterize biological surfaces with sub-micron precision, but also determine the elastic behavior of samples with nanoindentation-based approaches (Qian and Zhao 2018). The use of AFM for the study of collagens has allowed the exploration of its ultrastructure properties (Strange et al. 2017), including recent investigations of dentinal collagen from primary teeth in health and disease (Ibrahim et al. 2019). However, the accumulation of MGO-derived AGEs in the organic matrix of dentin associated to ageing, as well as the impact of these AGEs on nanomechanical changes at the single collagen fibril level have not yet been explored. Therefore, the aim of this work was to characterize the accumulation of MGO-based AGEs in the organic matrix of human dentin as a function of aging and associate these changes with alterations in the nanomechanical and ultrastructural properties of dentinal collagen.

## Materials and methods

### Tooth sample collection

Ethical approval was obtained from the local University Ethics Committee at Pontificia Universidad Católica de Chile (#180426002). Sample collection was carried out at the School of Dentistry Clinical Center (CODUC) in Santiago, Chile, following written informed consent. Twelve caries- and restoration-free teeth, extracted due to orthodontic or rehabilitation treatments from systemically healthy individuals were employed for this study. These samples were empirically divided into two different age groups (<26 year-old, and >50 year-old, n=6 per group) to characterize aging-related changes. Upon collection, teeth were washed in phosphate buffer saline (PBS, 1x), mechanically debrided to remove all remnants of periodontal tissue and dental pulp, and stored in 70% ethanol solution for 72 hours. Samples were then washed 3x in PBS and stored in PBS at 4°C before sectioning.

### Dentin section preparation and surface demineralization

Following tooth debridement and washing, samples were fully embedded in a dental acrylic resin (Marche Acrylics, Chile) and sectioned utilizing a SP1600 hard tissue microtome (Leica Biosystems, US). From these sections, multiple dentin samples from the crown and root (above and below the cementoenamel junction, respectively) were obtained with an individual thickness of 200 µm (**Figure 1A**) and collected immediately for further processing. For surface characterization, dentinal sections were further demineralized by washing with a solution of 37% phosphoric acid (Sigma-Aldrich, US) for 30 sec, rinsed with a 5% sodium hypochlorite solution (Sigma-Aldrich, US) for 10 sec, and further rinsed 3x with ultrapure H_2_O (Fresenius, Chile) (**Figure 1A**) (Ibrahim et al. 2019). Finally, samples were softly airdried for 1 hour semi-covered and transferred immediately to the microscope for assessment.

**Figure 1:**
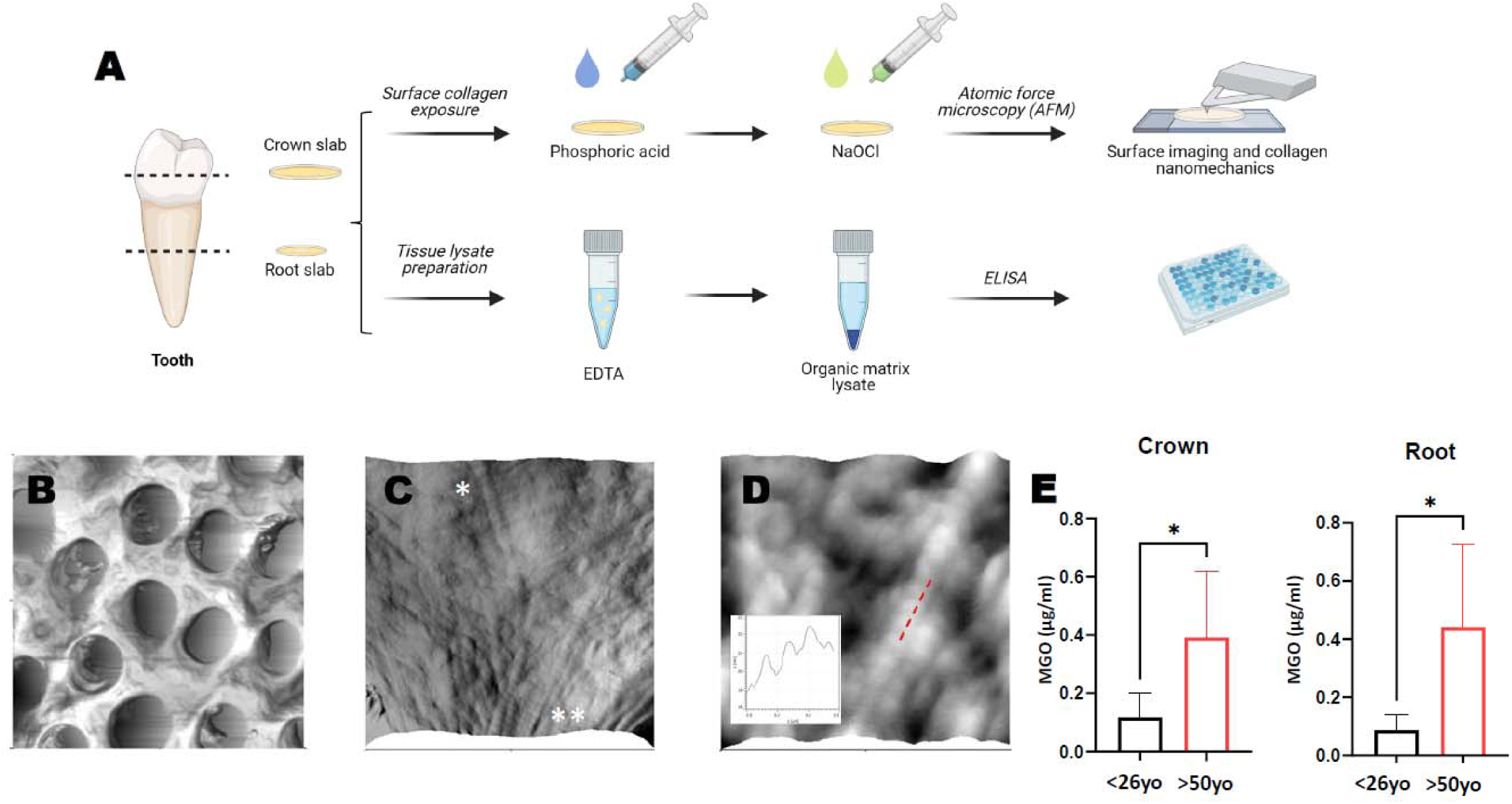
Study design and quantification of MGO-derived AGEs in human dentin. A) Overview of tooth sample preparation for atomic force microscopy (AFM) characterization and MGO-derived AGE quantification. Both crown and root dentinal samples from each tooth were processed for AFM and ELISA analysis. B) 20×20 µm AFM 3D reconstruction (from height channel) of crown dentin following surface demineralization protocol, displaying characteristic dentinal tubules. C) 2.5 × 2.5 µm AFM 3D reconstruction image of exposed intertubular (IT, *) and peritubular (PT, **) collagen fibrils. D) 1×1 µm AFM 3D reconstruction confirming the ultrastructure of collagen represented by regular d-banding periodicity (inset: collagen fibril profile from area market with red line). E) MGO-derived AGE quantification from the organic matrix of demineralized dentin from crown and root of patients <26yo and >50yo, showing an increase in MGO content in older samples (n=6; mean ± SD; *p<0.05; *t-test*).

### Determination of methylglyoxal-derived AGE content in dentin by ELISA

For the preparation of dentin tissue lysates, dentin sections were crushed into 1.5 ml Eppendorf tubes with the aid of pellet pestles (Sigma, US) and demineralized in a solution of 0.5□M ethylenediaminetetraacetic acid (EDTA) in 50□mM TRIS buffer (pH 7.4) for 72□hours at 4°C on an orbital shaker (Shinno et al. 2016). Protein content was measured using BCA assay, and samples were adjusted to the same concentration in EDTA TRIS buffer and diluted to 100µg/ml with 0.1% BSA in PBS.

The presence of MGO-derived protein adducts was measured in the organic matrix of dentin by using a competitive ELISA (Abcam, UK) according to manufacturer’s instruction. Briefly, protein binding 96-well plates were coated with 100 µl 500 ng/ml MGO conjugate in PBS and incubated over night at 4°C. Plates were then washed with assay wash buffer, blocked with assay diluent for 1 hour at room temperature, and washed again. Wells were incubated with sample or assay standard (50 µl) for 10 min, and subsequently 1x anti-MGO antibody (50µl) was added. After 1 hour incubation on an orbital shaker at room temperature, wells were washed and incubated with secondary antibody (HRP-conjugated). Finally, the assay was developed with substrate solution (R&D Systems) for 20 min, and the reaction was stopped with 1M H_2_SO_4_ and analyzed with a Tecan Sunrise microplate reader (Tecan, Austria).

### Atomic force microscopy (AFM) imaging and nanomechanical characterization of dentinal collagen

For this investigation, all AFM experiments were carried out with an Asylum MFP 3D-SA (Asylum Research, US) utilizing TAP300GD-G cantilevers (k∼24 N/m; BudgetSensors, Bulgaria). Dentinal samples were attached to magnetic microscopy discs with double-sided tape, and both height and amplitude channel images of substrates were obtained in intermittent contact mode in air. Each cantilever was calibrated before experimentation utilizing proprietary software, and a minimum of three 5×5 µm, 256×256 pixel scans were obtained on representative areas of each sample. Following topographical imaging, force-distance curves were obtained on individualized collagen fibrils by employing an adaptation of a previously published approach (Ibrahim et al. 2019). Briefly, collagen fibrils were indented with a maximum loading force of 30nN and a constant loading speed, along the center of each fibril in both the peri-tubular (PT) and inter-tubular (IR) dentin regions. A total of 100 force-distance curves were obtained per image (50 PT + 50 IT), for a total of 300 curves across the three areas of each sample. From the resulting force-distance curves, dentinal collagen Young’s moduli (YM) were calculated utilizing the Derjaguin, Muller, and Toporov (DMT) model in the proprietary Asylum Research AFM software (v. 16.10.211) (Schuh et al. 2021).

### Principal component analysis (PCA)

PCA analysis was carried out to further understand the association amongst all studied nanomechanical and molecular variables in both coronal and root dentin amongst all patients (Jolliffe and Cadima 2016). For this analysis, all values for MGO concentrations and collagen elasticity (in PT and IT dentin) for each patient were considered as independent variables. The PCA analysis were also performed on the <26yo and >50yo groups separately for better data visualization. From the molecular and elasticity variables, 5 principal components (PCs) were generated and selected based on the Kaiser rule (PCs with eigenvalues greater than 1.0). The loading plot (of the 1st and 2nd PCs) of each group was generated and compared for <26yo vs. >50yo and crown vs. root groups with Origin Pro 2018 (OriginLab Corporation, Northampton, MA). Score plots were obtained with GraphPad Prism 9 (GraphPad Software, San Diego, California USA).

### Statistical analysis

All data was tabulated and analyzed using the GraphPad Prism 9 software. After outlier detection, statistical significance was assessed with either linear regression, t-test, or Kruskal-Wallis tests, considering a significance value of p<0.05. Elasticity values are expressed as median with 95% confidence interval (95% CI).

## Results and discussion

Utilizing an adaptation of a previously published approach, it was possible to obtain dentinal sections from tooth samples from <26 year-old (n=6; 22 ± 2.1 years) and >50 year-old (n=6; 58 ± 6.9 years) patients, from which the organic matrix was exposed (Ibrahim et al. 2019) (**Figure 1B**). In all cases, the exposure of dentinal collagen in permanent teeth was found to be reproducible and to preserve the native appearance of fibrillar collagen, displaying its characteristic morphological markers including D-banding periodicity under the AFM (**Figure 1C and 1D**) as previously observed (Habelitz et al. 2002; Fawzy et al. 2012).

To determine the age-dependent accumulation of MGO-derived AGEs in the organic matrix of crown and root dentin, dentinal sections were demineralized with an EDTA solution and assessed with competitive ELISA (**Figure 1A**). In both crown and root dentin, we found a significant increase of MGO-derived AGEs in the >50yo group compared to <26yo patients (**Figure 1E**; p<0.05). Previous research has shown that MGO-derived AGEs accumulate in both the dental pulp and periodontal tissues (Retamal et al. 2016; Delle Monache et al. 2021), thus it is likely that perfusion from these tissues may promote MGO formation in dentin over time. Unlike bone, dentin has no turnover which facilitates the accumulation of AGEs in its organic matrix, similar to other slow turnover tissues such as tendons (Birch 2018). Levels of pentosidine in teeth have been observed to increase with aging associated to mechanical alterations (Shinno et al. 2016; Greis et al. 2018), and the AGE N-carboxymethyl lysine was also found to accumulate in dentinal collagen and increase mechanical resistance (Miura et al. 2014). Despite studies reporting the presence of amorphous proteins (Ravindran and George 2015), the organic matrix of dentin is predominantly comprised by collagen (specifically type-I collagen) (Goldberg 2011), which functions as the main target for MGO modification.

Following MGO-derived AGE quantification, a topographical characterization of the dentinal collagen matrix was performed on tooth crown samples with intermittent contact AFM. Imaging across all six <26yo samples demonstrated the presence of collagen fibrils throughout both the PT and IT regions with a clear D-banding register (**Figure 2A**). However, >50yo samples showed reduced presence of collagen fibrils and an increase in areas of wrinkling and loss of register (**Figure 2A**, asterisks), known to be representative of collagen aging in human tissues (Ahmed et al. 2017). Subsequently, nanomechanical analysis of PT collagen fibrils in the <26yo group showed YM mostly in the <10 GPa range (**Figure 2B**), with an overall median of 6.43 (95% CI[6.25-6.77]) GPa across all six samples. In the >50yo group however, there is a significant increase in PT collagen elasticity to a median of 9.25 (95% CI[8.73-9.73]) GPa (**Figure 2C**; p<0.0001, Kruskal-Wallis test). AGE accumulation has been shown to increase the YM of collagen in both in-vitro and tissue experiments (Verzijl et al. 2002; Depalle et al. 2015; Panwar et al. 2015; Schuh et al. 2021), suggesting that the increase in elasticity observed in PT collagen is mediated by the accumulation of MGO adducts and other potential AGEs and crosslinks (Lederer and Bühler 1999). As PT dentin is located immediately adjacent to the dentinal tubules, the presence of glucose and MGO in the pulp-derived perfusing fluid could explain collagen modification and the formation of AGEs over time (Miura et al. 2014).

**Figure 2:**
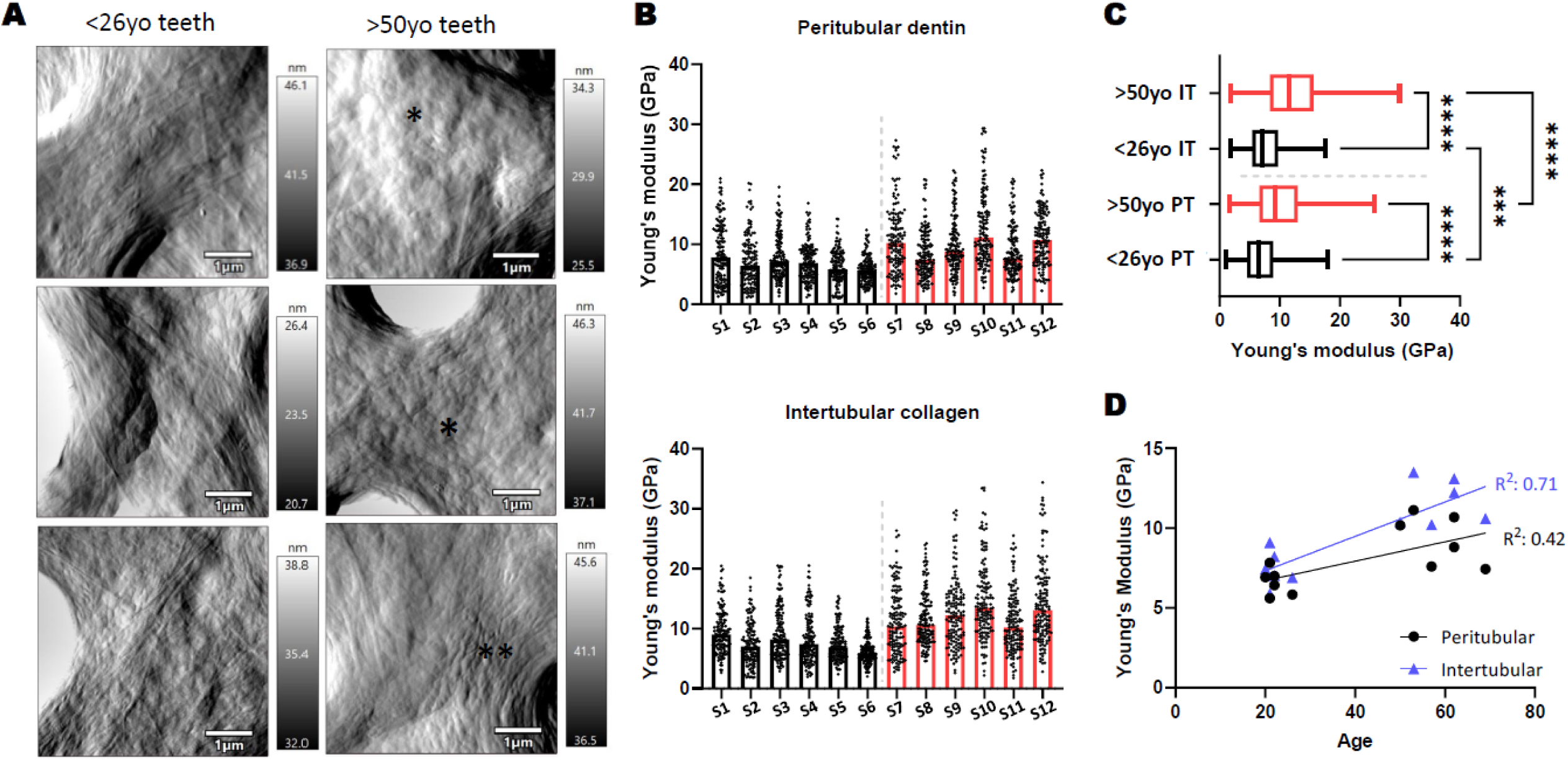
Ultrastructural and nanomechanical characterization of peritubular (PT) and intertubular (IT) collagen in coronal human dentin. A) Representative AFM amplitude images of the dentinal organic matrix in young and aged teeth, displaying both PT and IT collagen (* and ** indicate areas with loss of fibril register and fibril wrinkling, respectively). B) Collagen fibril YM for each examined tooth in both the PT and IT regions (n=150 force-distance curves per region, bars represent medians). C) Pooled YM collagen elasticity values for the <26yo and >50yo groups in both PT and IT regions (***, p<0.001; ****, p<0.0001; *Kruskal-Wallis test*). D) Linear regression plotted for tooth age vs YM for both PT and IT collagen fibrils.

Regarding IT collagen, it was also found to have higher YM than PT collagen in both age groups (**Figure 2C**; p<0.0001, Kruskal-Wallis test), and the mechanical properties of IT collagen were similarly found to vary as a function of age (7.09 (95% CI[6.86-7.39]) GPa and 11.59 (95% CI[11.2-12.06]) GPa for the <26yo and >50 yo groups, respectively) (**Figure 2B**). Overall, the age-associated increase in elasticity follows a linear trend for both PT (p<0.05) and IT collagen (p<0.001) (**Figure 2D**). This data suggests that AGE accumulation in the collagen matrix of dentin is not exclusive to the PT region, as IT collagen also increases its YM as a function of age. The <26yo group displays a very homogeneous grouping of YM for both PT (interquartile range, IQR=4.35) and IT (IQR=4.27), which spreads across a wider range of values in the >50yo group (IQR=6.37 and 7.24 for PT and IT, respectively). This observation suggests that despite an overall increase in YM, there are important individual variations regarding the aging process of dentinal collagen amongst patients (**Figure 2B**).

Following the characterization of crown dentinal collagen, root sections were also prepared and analyzed by AFM with the same approach (**Figure 1A**). Similar to crown dentin, fibrillar collagen was clearly observed in root dentin from <26yo samples and organized in both PT and IT areas, whereas the ultrastructure of collagen in the >50yo group included diverse areas with lack of register and loss of clear fibril morphology (**Figure 3A**). These morphological changed were also accompanied by an increase in YM for both PT and IT in the >50yo group compared to the <26yo group (**Figure 3B**). The median PT collagen elasticity values increased from 6.57 (95% CI[6.3-6.83]) GPa to 8.95 (95% CI[8.53-9.27]) GPa, and median IT values increased from 8.25 (95% CI[7.99-8.54]) GPa to 9.87 (95% CI[9.47-10.37]) GPa (**Figure 3C**; p<0.0001, Kruskal-Wallis test). These nanomechanical values of tooth collagen are similar to those previously reported by Ho et al. in demineralized cementum (4-7GPa) (Ho et al. 2007). Also, similar to crown dentin, the elasticity of PT collagen was found to increase linearly with age (p<0.001); however, the same association was not found for IT collagen, as it displayed higher heterogenicity in both groups (**Figure 3D**). Overall, our results suggest that mechanical changes in collagen associated to aging are widespread throughout dentin, involving both PT and IT collagen in crown and root.

**Figure 3:**
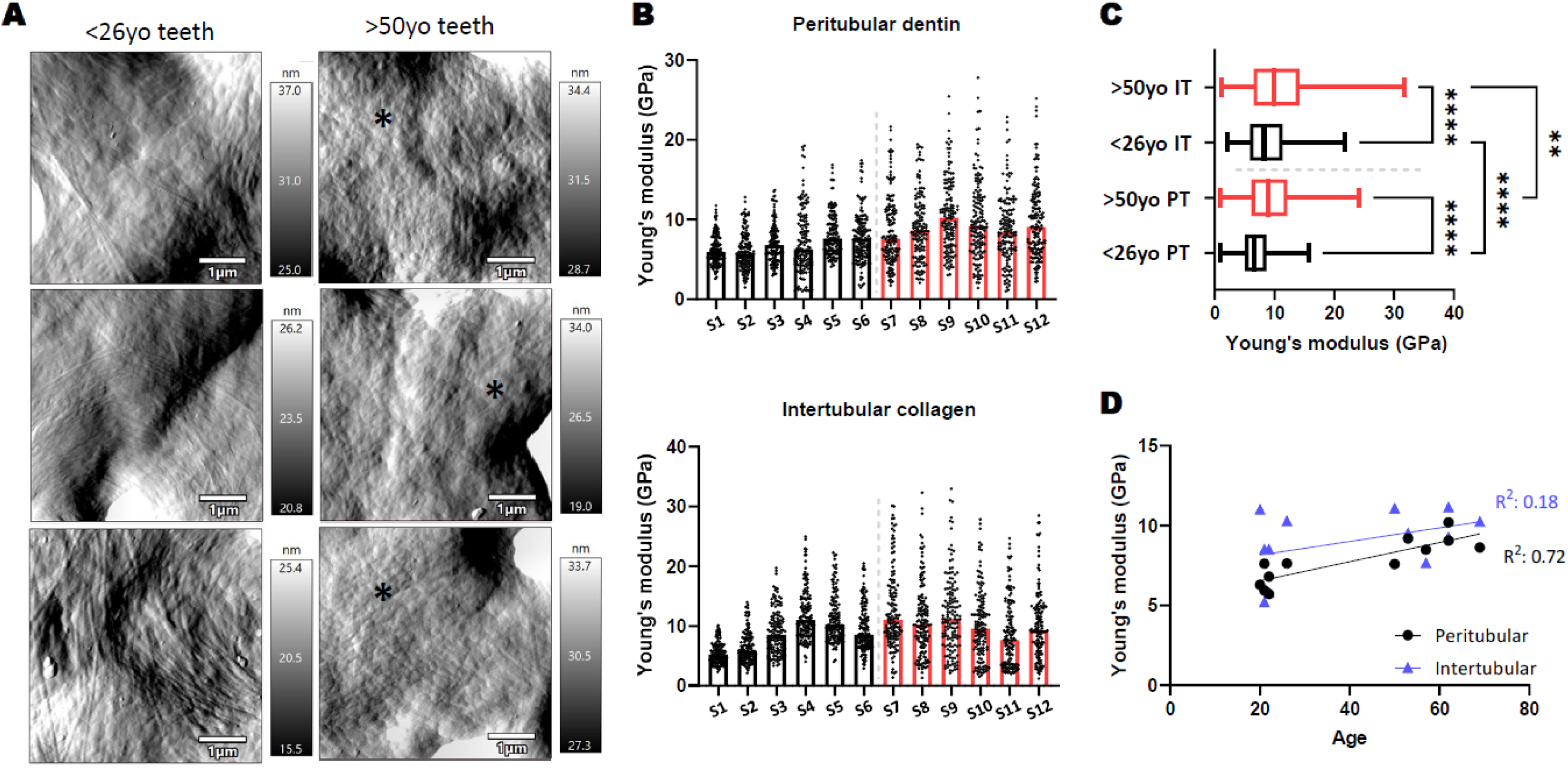
Ultrastructural and nanomechanical characterization of peritubular (PT) and intertubular (IT) collagen in radicular human dentin. A) Representative AFM amplitude images of the dentinal organic matrix in young and aged teeth, displaying both PT and IT collagen (* indicates areas with loss of fibril register). B) Collagen fibril YM for each examined tooth in both the PT and IT regions (n=150 force-distance curves per region, bars represent medians). C) Pooled YM collagen elasticity values for the <26yo and >50yo groups in both PT and IT regions of root dentin slabs (**, p<0.01; ****, p<0.0001; *Kruskal-Wallis test*). D) Linear regression plotted for tooth age vs YM for both PT and IT collagen fibrils in the root portion of each tooth.

As the final step, a PCA analysis was carried out to further understand the association amongst all studied nanomechanical and molecular variables in both coronal and root dentin. PCA is a statistical method that reduces a determined number of intercorrelated variables into dimensionless parameters called principal components (Jolliffe and Cadima 2016), and has been recently used in dentistry to facilitate the interpretation of complex datasets (da Fontoura et al. 2015; Elbahary et al. 2020). Overall, when MGO levels and IT and PT collagen nanomechanical variables were analyzed together, samples showed clear clustering in two distinct groups representing <26 yo and >50 yo patients, distributed along the PC1 axis, with no clear influence of patient gender on these groupings (**Figure 4A**). The intra-group variability amongst the <26 yo samples (blue dashed line) was reduced compared to the >50 yo (green dashed line), suggesting that the accumulation of age-related markers in dentinal collagen over time is not uniform amongst all individuals.

**Figure 4:**
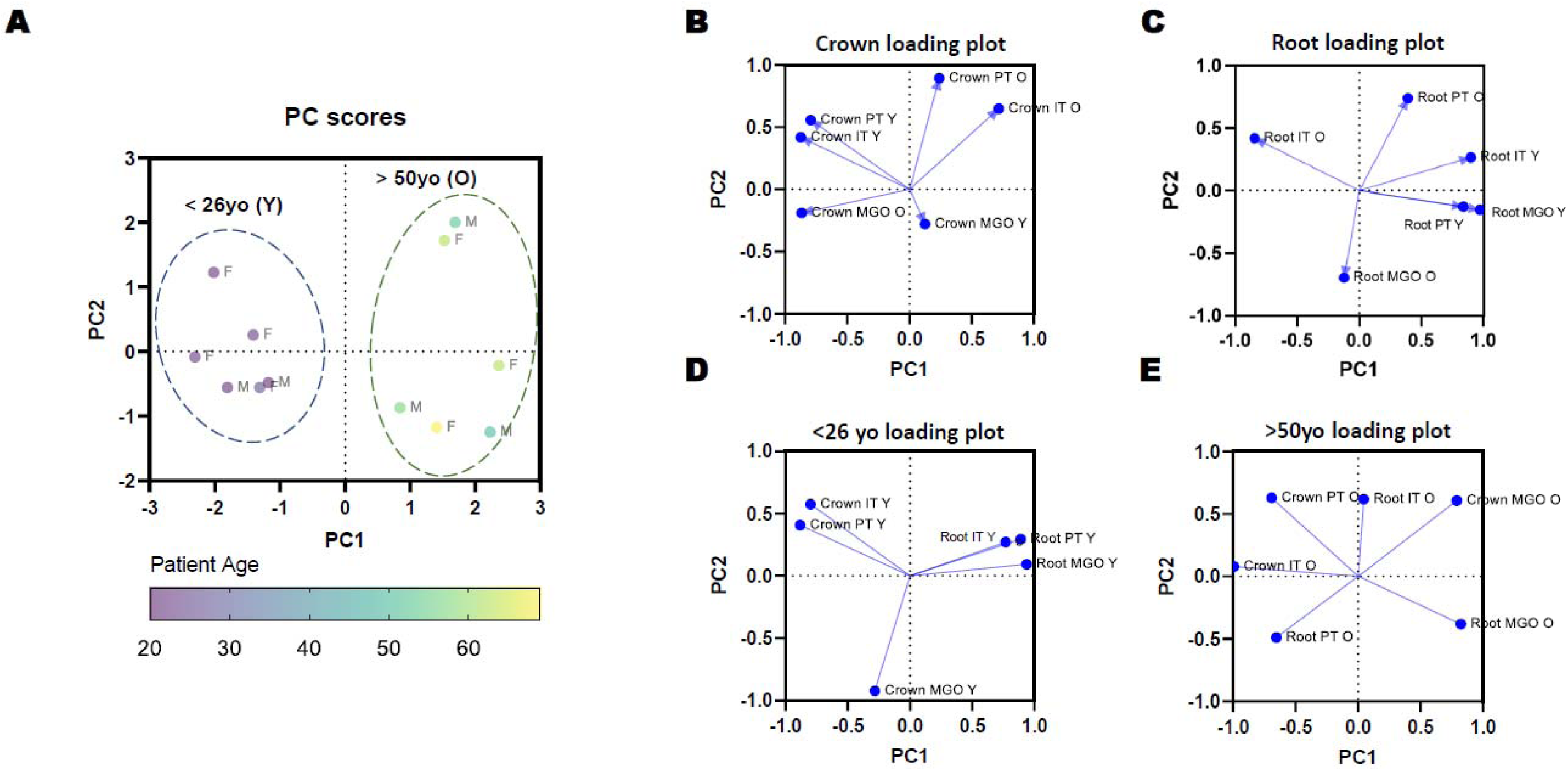
Principal component analysis (PCA) of molecular and nanomechanical aging parameters in human dentin. A) Score plot of the PCA for all studied variables, demonstrating clustering of samples along the PC1 as a function of patient age in the <26yo (younger patients, Y) and >50yo (older patients, O) groups. B) and C) Loading plots describing the association of all studied variables in both crown and root dentin, respectively. D) and E) loading plots describing the relationship of aging variables in the <26yo and >50yo groups, respectively.

Subsequently, specific PCA analyses by location and age group were performed (**Figure 4B-E**). In the crown, the mechanical properties of both PT and IT collagen were found to differ greatly between the <26 yo and >50 yo groups (**Figure 4B**). A similar age-associated difference was observed for MGO-adduct accumulation in the crown, suggesting that nanomechanical changes and MGO accumulation in the organic matrix of coronal dentin are important markers for tooth ageing. In the root, the strongest difference amongst the <26 yo and >50 yo groups was given by the mechanical properties of IT collagen, which separated mostly along the PC1 axis (**Figure 4C**). Thus, the variances observed among these two PCA loading plots suggest different “aging profiles” for both crown and root dentinal collagen, which could potentially have important therapeutic implications in restorative and adhesive dentistry approaches. Furthermore, PCA was also utilized to obtain profiles for the <26yo group (**Figure 4D**) and <50yo group (**Figure 4E**) samples. In younger patients, there are no clear differences between the mechanical properties of PT and IT collagen; however, the YM of collagen vary importantly between crown and root (**Figure 4D**). A different pattern is observed in >50yo patients, where there is less association amongst the studied variables, and separation is explained by a combination of the PC1 and PC2 components (**Figure 4E**). These distinctive nanomechanical and molecular profiles observed in >50yo samples strengthen the importance of further understanding how the process of aging impacts the mechanical properties of dentinal collagen Additionally, future work is needed to understand the potential impact of these aging-associated collagen changes on restorative dentistry approaches such as adhesive bonding to dentin and hybrid layer formation (Breschi et al. 2018), on bacterial adhesion and degradation during dentinal and root caries progression (Schuh et al. 2021; Álvarez et al. 2021), and on root caries restoration in the elderly.

## Conclusions

A reproducible *ex-vivo* model has been utilized to effectively characterize the ultrastructural and nanomechanical aging-associated changes of resident collagen fibrils within the organic matrix of human dentin. An increase in dentinal MGO was observed in >50yo compared to <26yo patients, in both crown and root. AFM nanoscale characterization demonstrated morphological alterations in the organic matrix of >50yo crown dentin samples, including loss of collagen ultrastructure and fibril banding. Furthermore, higher YM values in both PT and IT collagen fibrils were found in the >50yo group that increased linearly with age. Similarly, root-derived dentinal samples also displayed morphological changes and an increase in collagen fibril elasticity in the >50yo group, which was more pronounced in PT collagen. Finally, PCA analysis showed the formation of two clusters amongst samples from similar age groups that was mostly determined by differences in PT and IT crown elasticity as well as MGO-derived crosslink formation in crown dentin. Within each age group, variability was mostly given by differences in the elastic behavior of IT collagen in root dentin. Overall, these results further demonstrate that the human dentinal collagen matrix undergoes important age-related nanomechanical changes, with a potential impact on restorative dentistry approaches and dentinal biofilm formation in the elderly.

## Data Availability

All data is reported in the manuscript and are available from the corresponding author on request and review.

## Acknowledgements

This work was supported by the ANID FONDECYT Iniciación Grant #11180101 and Millennium Science Initiative #P10-035F. Figure 1A was created with Biorender.com. Authors would like to thank Camila Ramos for her support with sample collection.

